# Strategy-based Cognitive Rehabilitation for Childhood Brain Tumour: Protocol for an Acceptability and Feasibility Trial of the Fatigue, Learning, and Memory Enrichment (FLaME) intervention

**DOI:** 10.1101/2025.02.08.25321926

**Authors:** Charlotte P. Malcolm, Gerard Anderson, Victoria King, Deborah Ridout, Daniel Stark, Sara Shavel-Jessop, Emily Bennett, Antony Michalski, Tara Murphy, Faraneh Vargha-Khadem

## Abstract

**Background:** Medical treatments have improved survival rates for paediatric brain tumour (PBT), but the condition and treatment continue to be associated with significant cognitive morbidity. Nearly all survivors will experience some degree of cognitive impairment (neurocognitive ‘late effects’) that has a cascading impact on the development of intellectual and academic skills, quality of life, mental health, vocational attainment, and functional independence. Longstanding cognitive fatigue is also a prevalent symptom for survivors of PBT and further impacts engagement with therapeutic interventions and quality of life. Cognitive rehabilitation is recommended in national healthcare guidance and frequently requested by patients and families but rarely implemented due to a limited evidence base and poor feasibility and acceptability. There are currently no therapeutic interventions for cognitive fatigue for PBT survivors.

**Aims & Objectives:** We aim to establish feasibility, acceptability, and preliminary efficacy for strategy-based cognitive rehabilitation for PBT. The study will determine if there is benefit to feasibility and acceptability when cognitive fatigue management is integrated to cognitive rehabilitation.

**Methods:** Thirty-six 7-17 years olds diagnosed with PBT will be recruited from Great Ormond Street Hospital. Participants will be randomised to either 1) a 12-week cognitive rehabilitation intervention with integrated cognitive fatigue management, 2) a 6-week cognitive rehabilitation intervention alone, or 3) standard care. All participants will have received neuropsychological assessment identifying difficulties with cognition and fatigue. Feasibility (e.g., attrition, retention, adherence) will be assessed through the trial. Acceptability will be measured throughout using questionnaires and interviews based on the Theoretical Framework for Acceptability and satisfaction rating scale. Preliminary effectiveness data will be gathered pre- and post-intervention using standardised measures of cognitive skills, fatigue, quality of life, % school attendance, and goal-based outcomes.

**Outcome:** The findings will be used to determine the appropriate rehabilitation intervention for a larger, multicentre randomised controlled trial.

**PLAIN ENGLISH SUMMARY:** Medical treatments have improved survival rates for children with brain tumours. However, most children experience long-term difficulties with ‘cognition’ (thinking skills such as memory and paying attention) and cognitive fatigue (excessive mental tiredness) after treatment. Thinking difficulties and fatigue can affect a child’s ability to learn, and their social and emotional wellbeing. National guidance recommends treatment called ’cognitive rehabilitation’ which teaches skills to improve or manage cognitive difficulties. Families often request this, but it is not usually available due to little research. Fatigue may also get in the way of children using and benefiting from cognitive rehabilitation. No research study yet has offered a fatigue treatment for children recovering from brain tumours. The study aims to see if it is practical and helpful to families to provide cognitive rehabilitation for children affected by brain tumours. The treatment focuses on strategies to help cognition. We will see if adding strategies to manage fatigue helps. We will include thirty-six 7–17-year-olds who have been cared for at Great Ormond Street Hospital for brain tumour. All participants will have had an assessment describing cognitive strengths and weaknesses as part of usual care. Participants will be randomly allocated to one of three groups: 1) cognitive rehabilitation with fatigue management (12 weeks), 2) cognitive rehabilitation only (6 weeks), or 3) usual care. Each child and their carer will complete questionnaires before, during, and after the treatment, and an interview at the end of the treatment. This information will help the researchers see if families find the treatment helpful and practical to take part in, and if adding fatigue strategies is beneficial. Researchers will look at information such as the number of appointments attended, feedback about the treatment, and information about fatigue levels, cognition, and wellbeing. The findings will be used to develop a UK-wide study.

## 1. INTRODUCTION

### 1.1 Background & Rationale

Treatment advances for paediatric brain tumour (PBT) in recent decades have substantially improved mortality rates but come at significant cost in cognitive morbidity alongside the long-term effects of the tumour itself. Up to 100% of children treated for PBT experience some degree of cognitive difficulty despite most having achieved typical cognitive development prior to diagnosis (1). Most children experience a constellation of long-term cognitive impairments that impact quality of life, mental health, access to education, academic and vocational attainment, and functional independence in adulthood. The most common neurocognitive impairments are in processing speed, attention, working and long-term memory, and visual-motor function which increase with time since treatment and result in the insidious secondary slowing of intellectual and academic progress over time (neurocognitive ‘late effects’), with an average loss of 18 Full-Scale IQ points by early adulthood (2, 3).

The well documented neurocognitive late effects inform the strong emphasis on neurorehabilitation in national guidance for PBT (4, 5). Interventions aimed to alleviate the burden of cognitive impairment are paramount but rarely implemented (6). Cognitive rehabilitation typically involves systematic interventions to restore and/or compensate for cognitive impairment (7) including 1) massed practice on computerised cognitive training tasks (drill-based practice, e.g., Cogmed), 2) internal and metacognitive strategy-use to optimise cognition (e.g., generalisable elaborative encoding techniques such as mnemonics), and/or 3) external compensatory aids (e.g., visual reminders). Cognitive rehabilitation is almost entirely unavailable for PBT (8) with a paucity of research and poor feasibility and acceptability of trialled interventions. Drill-based computerised rehabilitation has been the focus of the small number of available studies, however, the feasibility and acceptability of this approach for PBT is low (9) and has well documented problems in poor generalisation to skills beyond the trained task (e.g., academic skills) and poor maintenance of improvements (10). The focus of drill-based interventions is typically to remediate specific cognitive deficits in isolation which neglects the broader profile of multiple co-occurring neuropsychological difficulties common after PBT.

Rehabilitation in paediatric acquired brain injury has greatest potential when it includes internal and metacognitive strategy-use, is adapted for developmental level, and delivered within a systemic context (7). There is currently one good quality randomised controlled trial (RCT) of a cognitive rehabilitation intervention that incorporates strategy-use for PBT (11). The intervention resulted in improvements in parental report of child inattention and academic attainments, albeit with small effect sizes. The program involved strategy-use training, but also included a demanding drill-based practice component (requiring 40 hours of practice), with only 60% of participants completing the intervention. A subsequent study (12) omitted the drill-based practice and extended the strategy-use component (15 sessions, 20 hours in total), combining it with systemic support in a smaller pilot study. The power and generalisability of the findings are limited by a small sample size, however significant improvement for written expression was found along with trends towards improvement on neurocognitive measures. Participants also rated high levels of satisfaction, particularly because the intervention improved their understanding of their cognitive strengths and weakness.

Despite some promising findings for strategy-based cognitive rehabilitation, poor feasibility is reflected in low completion rates. Cognitive fatigue (extreme mental tiredness) is one of the most frequently reported and distressing chronic symptoms after PBT (13). Cognitive fatigue can impact adherence to interventions, as well as participation in settings where children make most use of rehabilitation strategies such as in education and activities of daily living. Despite cognitive fatigue being both common and impairing, it has been significantly overlooked in both research and clinical interventions for PBT. No cognitive rehabilitation intervention for PBT has addressed the high prevalence of cognitive fatigue that could predictably limit engagement and completion of rehabilitation.

There have been no published trials to date of cognitive rehabilitation interventions for child survivors of brain tumour in the United Kingdom and within the National Health Service (NHS). The Department of Health and Social Care ‘Brain tumour research: task and finish working group report’ (14) strongly advocates that research should focus on quality of life, including “living with the long-term cognitive effects of surgery and radiotherapy” (pg.21); a priority that should be “embraced by the research community and funders” (pg. 21). The proposed research study addresses this public health priority by trialling the acceptability and feasibility of an intervention to address the long-term cognitive effects of brain tumour and treatment that impact on multiple facets of survivorship and quality of life. The study will 1) assess the acceptability and feasibility of a novel cognitive rehabilitation intervention for PBT, and 2) determine whether incorporating cognitive fatigue management improves feasibility, acceptability, and patient-reported benefit.

### 1.2 Theoretical approach to acceptability

Acceptability will be evaluated using the Theoretical Framework for Acceptability (TFA (15, 16)). The TFA offers a robust framework for evaluating theoretically informed components of acceptability throughout and after an intervention by those who receive and deliver it. Examples of component constructs include affective attitude (the individual’s feelings about the intervention), burden (how effortful it is to participate), and perceived effectiveness (how the individual perceives the intervention to have achieved its goal).

### 1.3 Theoretical approach to outcome measurement

A major aim of cognitive rehabilitation is to improve everyday functioning and quality of life. Prevailing drill-based rehabilitation seeks to achieve this through remediating specific cognitive impairment. This approach naturally lends itself to selecting performance-based neuropsychological tests as the primary outcome measure. Impairment on performance-based tests is typically identified at pre-intervention and the test is repeated post-intervention to measure the extent of remediation. This approach has been found to yield either non-significant results or small effect sizes on measures other than those very similar to the drilled task (10), and practice effects from repeated cognitive testing can confound study results (11). Strategy-based cognitive rehabilitation shifts the focus of rehabilitation away from remediating impairment to developing metacognitive and compensatory mechanisms to better manage the impact of the cognitive impairment on everyday life. Studies focused on strategy-based rehabilitation in paediatric brain injury therefore have included outcome measures related to everyday function. On recent systematic review (7), strategy-based rehabilitation predictably does not produce change on performance-based cognitive tests. However, these studies demonstrate significant and powerful improvement on functional measures such as everyday executive function skills, problem-solving, goal-directed behaviour, daily living skills, individual rehabilitation goals, and quality of life. The findings have informed the focus of outcome measurement in the current study.

### 1.4 Study Objectives

The overarching study aim is to establish feasibility and acceptability for a strategy-based cognitive rehabilitation intervention for PBT, and any benefit to feasibility and acceptability by integrating cognitive fatigue management. The findings will be used to determine whether the cognitive rehabilitation intervention alone, or the same intervention with integrated fatigue management should be taken forwards to a definitive RCT.

Objectives include:

1. To assess whether the proposed cognitive rehabilitation intervention design is feasible to implement and acceptable to patients.
2. To assess whether feasibility and acceptability differ when fatigue management is incorporated to determine which intervention arm to take forwards to the definitive RCT.
3. To measure preliminary/limited-efficacy patient reported benefit and outcomes of the intervention arms relative to standard care.
4. To identify the optimal outcome measures for a larger scale RCT.
5. To document any barriers to recruiting a representative sample and acceptability of randomisation.
6. To identify any practical barriers to conducting an RCT intervention at the designated NHS site.

## 2. METHODS

### 2.1 Study Design and Setting

The feasibility study employs a randomised, parallel arm design with a standard care control group (see Figure 1: Study Flow chart).

**Figure 1:**
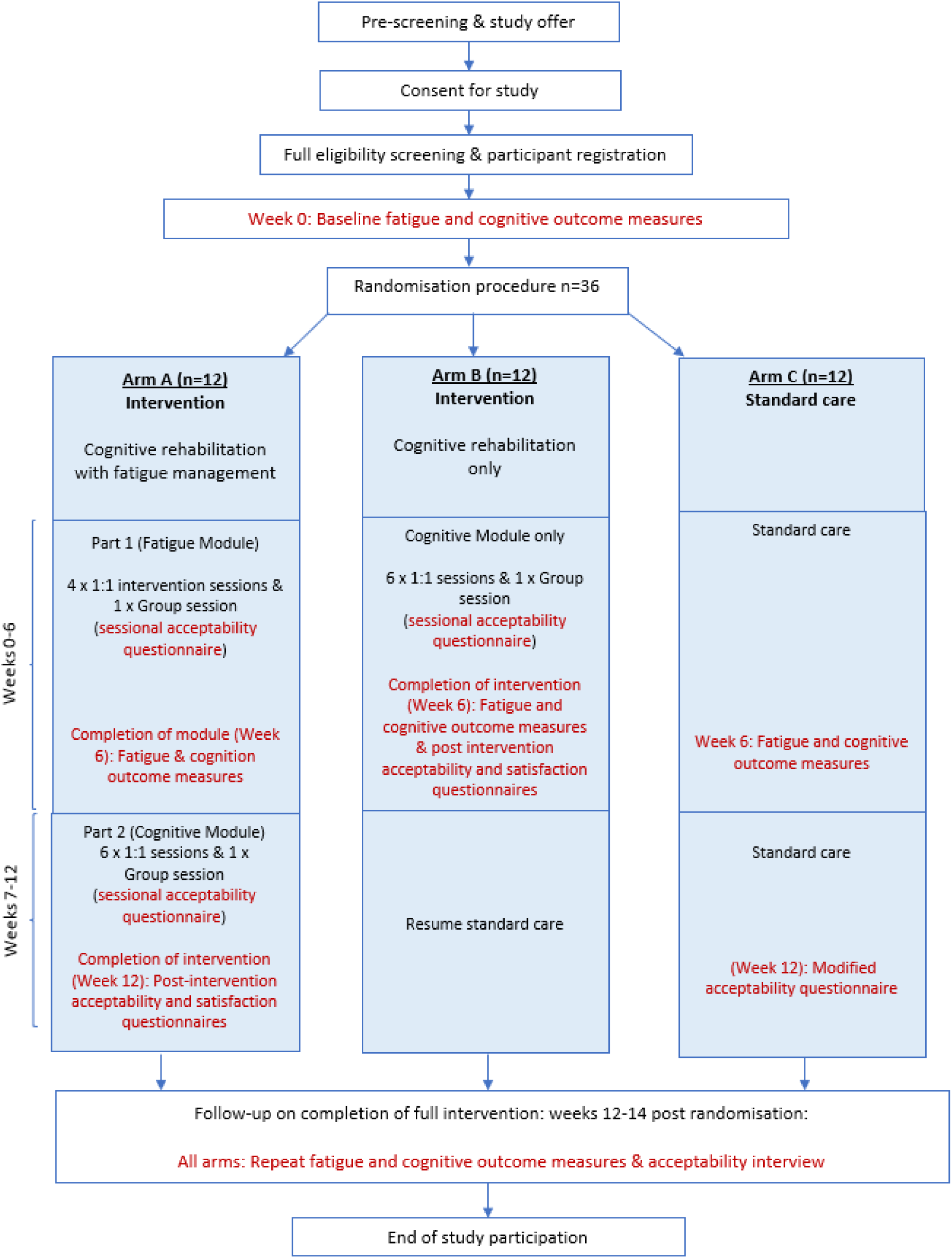
Study Flow Chart.

Thirty-six participants will be recruited and randomised to either 1) a 12-week block of intervention (cognitive rehabilitation with fatigue management), 2) a 6-week block of intervention (cognitive rehabilitation alone), or 3) 12 weeks of standard care. Randomisation will be undertaken centrally by the study team using GraphPad random assignment. Participants will be young people aged 7 years to 17 years, 11 months who have been diagnosed with a childhood central nervous system (CNS; brain) tumour, or received oncology treatment to the brain, and are current patients at Great Ormond Street Hospital for Children (GOSH). The study is a single-site study. All participants will be recruited at GOSH, with each participant completing a 14-week study period. All study procedures will take place at GOSH.

### 2.2 Eligibility Criteria

Potential participants will be invited to take part based on the following inclusion and exclusion criteria:

Inclusion criteria

1. Age range: 7 years to 17 years, 11 months.
2. Received diagnosis and/or treatment/surveillance at GOSH for a childhood tumour that involved the CNS (brain) and/or oncology treatment to brain.
3. Received or receiving a neuropsychological assessment/consultation at GOSH over the course of the study period or in the 48 months prior to the study period, or under active surveillance with the neuro-oncology multidisciplinary team during the study period.
4. At least 6 months post-diagnosis/acute treatment (surgery and/or radiotherapy), and 3 months post-return to school, with stable disease.
5. One or more scores outside of normal limits (i.e. 1 SD above or below the mean in the direction indicating difficulty) in at least one neuropsychological domain (on performance-based tests or questionnaire-based rating scales).
6. Report impairment (z-score > -0.67) in fatigue on one or more subscales of the PedsQL Multidimensional Fatigue Scale.
7. Capacity/competence of patient or parent/carer to provide informed consent.

Exclusion criteria

1. Completed or having another targeted formal psychological intervention for cognitive rehabilitation or fatigue in the past 6 months.
2. Sensorimotor (e.g., visual-motor) impairment only on neuropsychological assessment without additional cognitive difficulty.
3. Current substance misuse from self-report.
4. Currently receiving formal psychiatric care for a diagnosed mental health disorder (including active suicidal ideation), excluding ADHD treatment (if a child has a diagnosis of ADHD they should be treated).
5. Intellectual Disability based on a standard score of more than 2 standard deviations below the mean on a general adaptive behaviour composite and, where available, the General Ability Index of intellect.
6. Patient and parent/carer is unable to communicate verbally and in written form in English.

### 2.3 Intervention

The Fatigue, Learning, and Memory Enrichment (‘FLaME’) program was developed by members of the research team by incorporating strategies that have been trialled and found successful in fatigue (17–19) and cognitive rehabilitation (11, 12, 20–24) interventions for children. Strategies for cognitive fatigue include pacing, activity scheduling and monitoring, behavioural strategies to regulate stress, and basic psychoeducation (e.g., sleep hygiene). Strategies for cognition focus on regulating attention to information and optimising encoding and retrieval (e.g. chunking and elaborative semantic encoding strategies such as the PQRST method and mnemonics), age-appropriate problem-solving strategies, and compensatory strategies such as visual reminders. A paediatric neurocognitive interventions model (PNI model (20)) informed the theoretical development of the intervention. In the PNI model skills are targeted sequentially based on a developmental hierarchical model where foundational factors (e.g., psychosocial needs) are addressed first, followed by adult-supported compensatory strategies, with independent use of strategies for specific impairments delivered only once these earlier levels have been addressed. This approach was recently piloted to deliver strategy-based rehabilitation for memory impairment in children with paediatric traumatic brain injury, finding large effect sizes for improvements in everyday memory difficulties (25). Extending the PNI model for childhood brain tumour, cognitive fatigue is a common co-occurring difficulty that plausibly benefits from intervention at the foundation stage before subsequent levels of cognitive rehabilitation are delivered.

All interventional components are delivered by a Psychologist under the supervision of a senior Clinical Psychologist & Neuropsychologist. The full intervention is delivered over 12 weeks, inclusive of 10, 1-hour individual sessions (child and parent attends) and a separate parent and peer psychoeducational group session during each block (i.e. during the fatigue and cognitive intervention blocks). The group sessions reinforce the same psychoeducational components of individual sessions and are primarily to facilitate peer support. The cognitive rehabilitation-only intervention is delivered in a 6-week block consisting of 6 individual sessions (child and parent attends) and a parent and peer group session. Psychoeducational webinars for each block of cognitive fatigue and cognitive rehabilitation are provided to the child’s teacher/Special Educational Needs Co-ordinator (SENCo) to enhance systemic support and maximise generalisation of strategies. The intervention is delivered hierarchically with cognitive fatigue management introduced first (in the full intervention), followed by cognitive rehabilitation strategies with consideration to developmental stage of the child.

The intervention can be delivered in face-to-face or remote format depending on participant preference. Meta-analysis (26) and review (27) of similar remote skills-based neuropsychological interventions for young people with diverse neurological conditions have recently found remotely delivered interventions have high levels of feasibility and acceptability without compromise to fidelity and efficacy. They also have the advantage of greater geographical reach and access for families who may otherwise be unable to participate (12, 28). Criteria for discontinuing the intervention includes patient request, disease relapse requiring urgent medical treatment, and where the participant becomes ineligible based on the eligibility criteria.

### 2.4 Study data and outcome measures

#### 2.4.1 Demographic and clinical data

• Demographic data: age, date of most recent neuropsychological assessment, sex, ethnicity, living location, parental education and occupation.

• Clinical data: primary oncology diagnosis/tumour type, tumour location, tumour WHO grade, oncology treatment type (surgery, chemotherapy, and/or radiotherapy), co-morbid health conditions, diagnoses, and treatments.

#### 2.4.2 Neuropsychological assessment data

All children will have received protocol-based neuropsychological assessment/consultation within the clinical service prior to commencing the intervention. This includes standardised measures such as direct measures of intellectual function (e.g., Wechsler Intelligence Scale for Children – 5th Edition), attention (e.g., Conners Continuous Performance Task – 3d Edition), memory (e.g., Children’s Memory Scale), and indirect measures of everyday skills (e.g., Adaptive Behaviour Assessment – 3rd Edition, Behaviour Rating Inventory of Executive Function – 2nd Edition). Specific measures are administered according to the age of the child and presenting needs. This takes place as part of standard care and will be used to characterise the cognitive needs of the participant sample.

#### 2.4.3 Feasibility measures

##### 2.4.3.1 Demand

1. Acceptance/refusal rates for potentially eligible participants.
2. Documentation of ineligibility and refusal reasons.
3. The completed number of sessions out of the total planned and attrition rate across the lifecycle of the study.
4. Record of any bias in dropout (i.e. by demographic and clinical characteristics).
5. Length of time to recruit participants within the study recruitment window.
6. A qualitative logbook of any unanticipated challenges or ‘lessons learned’.

##### 2.4.3.2 Implementation, Practicality, Adaptation, and Integration

1. Fidelity: Clinician and observer report of fidelity using a checklist for the content coverage of the specific intervention session.
2. Participant adherence: Adherence to intervention strategies through completion rate of a brief home learning task related to the individual session content of the week. Adherence will also be discussed at post-intervention interviews.
3. Estimated cost analysis of resource required for the RCT and/or sustainability in the organisation.
4. Feasibility of data collection time points as indicated by attrition, missing, and unusable data.
5. Documentation of obstacles to recruitment in the organisation.
6. Mode of intervention delivery.

#### 2.4.4 Acceptability measures

The study will triangulate qualitative and quantitative data through TFA-informed questionnaire (16) and qualitative interviews. The TFA-questionnaires were developed for the study using a published template (16) as a session-by-session and post-intervention measure for each trial arm. Questionnaires were adapted according to the age of the child. Semi-structured interview schedules were also developed for children, parents/carers, and therapists delivering the intervention using the same template and research (29). The questionnaires and interview guides were independently evaluated by two researchers with advanced experience in psychometrics for paediatric health. To assess the construct validity of each item a process of ‘back coding’ (29, 30) was employed. Each researcher was sent the questionnaire and interview schedule items in a random order with a list of TFA constructs and were required to match each question to the correct construct and rate their confidence on a five-point scale (1=not at all confident, 5 = completely confident). This process indicated strong construct validity where 95% of items were correctly matched to their construct with a high degree of certainty (average 5 of 5 for each construct across both researchers). The only items not correctly matched on one occasion and by one researcher were the ‘affective attitude’ and ‘general acceptability’ items which were interchanged. A second researcher rated certainty of 4 out of 5 for the ‘affective attitude’ and ‘general acceptability’ items of the questionnaires, where all other items yielded certainty of 5/5 across all questionnaires and interviews. On discussion with the researchers, it was agreed that these two items had a high degree of conceptual overlap in questionnaire format and would likely be highly correlated. A decision was made to eliminate the general acceptability item from the questionnaires which is an optional item (16). The item was retained for the interviews however where it was a more distinct and could be explored with prompt questions.

The following TFA-measures will be used:

1. Session-by-session TFA-Questionnaire for parents & child.
2. Post-intervention or standard care TFA-Questionnaire for parents & child.
3. TFA-informed qualitative interview with participants on experience and acceptability of intervention or standard care, and acceptability of randomisation.
4. Post-intervention TFA-Questionnaire with qualitative feedback for education staff.
5. Post-intervention feasibility and acceptability interview with the therapist delivering the intervention.

The Satisfaction Questionnaire after Cognitive Skills Training Interventions – parent (12) will be used as a comparison to another published study.

#### 2.4.5 Preliminary outcome measures

The inclusion of outcome measures in the feasibility trial is for ‘limited-efficacy testing’ to inform a future fully powered RCT and for descriptive analysis of patient benefit by assessing 1) the measures’ sensitivity to change in the PBT population within the context of a feasibility trial, and 2) to estimate effect sizes of the measures (31). Outcome measures are administered at baseline, week 6, and weeks 12-14 before acceptability interviews (see Figure 1: Study Flowchart):

1. Fatigue measures

a. Primary outcome measure: Goal Based Outcome (GBO) for management of cognitive fatigue - parent & child (32)
b. Secondary outcome measures: Multidimensional Fatigue Scale – parent and child (33), individual daily fatigue analogue scale (child), % school attendance (from school report)
2. Cognitive measures

a. Primary outcome measure: GBO for management of cognitive difficulty – parent and child (32).
b. Secondary outcome measures: Behaviour Rating Inventory of Executive Function – second edition (BRIEF-II) – parent and teacher, PedsQL Core + Brain Tumour Cognitive Problems module – child and parent (34), The Brief Illness Perceptions Questionnaire – child (35)

These outcomes measures were selected due to their psychometric properties, suitability to the aims of strategy-based cognitive rehabilitation, and precedence and sensitivity to detecting change after rehabilitation interventions (18, 25, 36–40).

### 2.5 Participant timeline

A schedule of participant procedures is shown in Table 1.

**Table 1:** Schedule of Participant Procedures.

Key: A = Arm A (Full intervention), B = Arm B (Cognitive rehabilitation only), C = Arm C (Standard care)
| Procedure | Screening | Baseline | Wk1 | Wk2 | Wk3 | Wk4 | Wk5 | Wk6 | Wk7 | Wk8 | Wk9 | Wk10 | Wk11 | Wk12 | 12-14 |
| --- | --- | --- | --- | --- | --- | --- | --- | --- | --- | --- | --- | --- | --- | --- | --- |
| Informed consent | A,B,C |  |  |  |  |  |  |  |  |  |  |  |  |  |  |
| Collect demographics | A,B,C |  |  |  |  |  |  |  |  |  |  |  |  |  |  |
| Collect clinical information | A,B,C |  |  |  |  |  |  |  |  |  |  |  |  |  |  |
| Full eligibility screen | A,B,C |  |  |  |  |  |  |  |  |  |  |  |  |  |  |
| GBO for fatigue |  | A,B,C | Arms A & B at each 1:1 intervention session |  |  |  |  | A,B,C | Arms A at each 1:1 intervention session |  |  |  |  |  | A,B,C |
| MDFS |  | A,B,C |  |  |  |  |  | A,B,C |  |  |  |  |  |  | A,B,C |
| Fatigue VAS |  | A,B,C | A,B | A,B | A,B | A,B | A,B | A,B,C | A | A | A | A | A | A | A,B,C |
| School attendance % |  | A,B,C |  |  |  |  |  |  |  |  |  |  |  |  | A,B,C |
| GBO for cognition |  | A,B,C | Arms A & B at each intervention session |  |  |  |  | A,B,C | Arms A at each intervention session |  |  |  |  |  | A,B,C |
| BRIEF II |  | A,B,C |  |  |  |  |  | A,B,C |  |  |  |  |  |  | A,B,C |
| PedsQL core & BT modules |  | A,B,C |  |  |  |  |  | A,B,C |  |  |  |  |  |  | A,B,C |
| BIPQ |  | A,B,C |  |  |  |  |  | A,B,C |  |  |  |  |  |  | A,B,C |
| Group intervention session |  |  | Arm A & B 1 x session |  |  |  |  |  | Arm A = 1 session |  |  |  |  |  |  |
| Individual intervention session |  |  | Arm A = 4 x sessions, Arm B = 6 sessions |  |  |  |  |  | Arm A = 6 sessions |  |  |  |  |  |  |
| TFA sessional measure |  |  | Arms A & B at each intervention |  |  |  |  |  | Arms A at each intervention session |  |  |  |  |  |  |
| TFA post intervention questionnaire |  |  |  |  |  |  |  | B |  |  |  |  |  |  | A/C |
| Satisfaction with Cognitive Skills Training Questionnaire |  |  |  |  |  |  |  | B |  |  |  |  |  |  | A |
| TFA interview |  |  |  |  |  |  |  |  |  |  |  |  |  |  | A,B,C |

GBO=Goal Based Outcome, MDFS=PedsQL Multidimensional Fatigue Scale, VAS=Visual analogue scale, BRIEF II= Behaviour Rating Inventory of Executive Function second edition, PedsQL= Pediatric Quality of Life Inventory, BT=Brain Tumour, BIPQ= Brief Illness Perceptions Questionnaire, TFA=Theoretical Framework for Acceptability

### 2.6 Data recording, storage, and access

A Case Report Form (CRF) will be assigned for all participants entering the study. The CRF will be depersonalised and the participant and CRF will be assigned an anonymised code (Patient Identification Number; PID). All data recorded will meet the standards set out by the Sponsor’s local policies. An electronic Study Site File will be maintained on a secured GOSH Trust drive which will only be accessible to members of the research team at GOSH. All study source data (e.g., from patient records and study measures) will be documented in the electronic CRF. No data fields will be left blank. Where there is missing data, this will be coded with a pre-specified key. All data from the CRF will be transferred to an electronic study file in depersonalised format when the participant completes the study. The CRFs and study file will be held in duplicate in a secure GOSH Trust drive and Trust approved encrypted storage device and only accessible to the GOSH site research team. Where data is transferred from source data to CRFs, and from the CRF to study file, this will be checked for accuracy by a second member of staff (named in the CRF). The Chief Investigator (CI) or delegated individual from the research team will perform random regular audit of CRFs and the study file for accuracy (checking a minimum of a randomly identified 15%).

### 2.7 Data processing and analyses

#### 2.7.1 Quantitative data

A detailed statistical analysis plan will be produced prior to data analyses. All demographic, clinical, and neuropsychological assessment data will be summarised by group. Quantitative feasibility data will also be summarised for the sample. Quantitative data from TFA-questionnaires will also be used to compare and rank individual intervention sessions, the overall intervention, and compare to acceptability measures in the standard care arm. Means and standard deviations (or alternative as appropriate) for the satisfaction questionnaire will be described according to intervention arm.

Statistical analyses will mainly focus on change detected in the outcome measures (e.g., GBO, PedsQL, BRIEF-II). Means and standard deviations (or equivalent) will be described for the outcome measures for each group at each time point (baseline, week 6, and weeks 12-14), and compared using repeated measures ANOVA to address these two aspects of limited efficacy testing. The focus of the statistical analyses will be to estimate effect sizes (e.g., partial eta squared, Cohen’s d, or non-parametric equivalents as needed) for each measure. Effect sizes will then be ranked across the outcome measures to determine which is most sensitive to change to inform the future clinical trial. The 95% confidence intervals for the effect sizes will also be considered.

#### 2.7.2 Qualitative

Qualitative data generated by feasibility outcomes will be grouped into themes. Frequency rates for each theme will be calculated if this is appropriate. It will be used to accompany description of quantitative feasibility data.

The primary qualitative data will be generated by the TFA-informed interviews. This data will be analysed using thematic analysis (41), an analytic method that explores patterns and themes in qualitative data. Themes will be analysed according to the TFA. The final analyses will be shared with two participants to provide a member check of final themes and discussed with the Patient and Public Involvement Advisory Group.

### 2.8 Sampling and sample size

Thirty-six participants in total will be recruited into the study via convenience sampling of the current and retrospective (past 48 months prior to study start date) patient pool from the Neuropsychology service. Participants will be recruited continually until the sample is reached. Continual recruitment involves offering the study to potential participants as they come through the clinical service systematically, and by searching the service database backwards systematically (in reverse order of assessments completed) over the past 48 months and applying the inclusion and exclusion criteria to invite potential participants. Priority will also be given in this order to patients who are either due to turn 18 years old, transition to another service, or would become ineligible for another reason within the next 6 months. All potential participants will be contacted initially by a member of their healthcare team via telephone, letter, patient messaging, or by discussion with their neuropsychology clinician on completion of their neuropsychological assessment. The GOSH neuro-oncology multidisciplinary team will be made aware of the study objectives, inclusion, and exclusion criteria and can inform potential participants of the study. Only trained and delegated members of the healthcare team can discuss the study in detail or provide Patient Information Sheets (PIS). The rationale for this sampling strategy is to maximise recruitment of a broad range of potential participants who meet the essential inclusion criteria to answer key questions of feasibility and acceptability in this population to inform the future RCT.

The justification for the sample size comes from the NIHR Research Design Service (RDS) London evidence for sample sizes for feasibility studies (https://www.rds-london.nihr.ac.uk/resources/justify-sample-size-for-a-feasibility-study/) and associated evidence for estimating a standard deviation to power the future definitive trial. There is no formal way to power a standard deviation estimate which is a measure of variability, therefore various rules of thumb have been developed that the sample size for a feasibility trial should be between 24-50 (42, 43). A sample size of 36 (12 participants per arm of the study) is further justified by the feasibility and the precision this provides in estimating the mean and the variance (42). This can then be used to calculate power for a more precise hypothesis for a definitive randomised controlled trial.

### 2.9 Recruitment

Pre-screening will include screening potential participants according to the inclusion and exclusion criteria 1) on completion of prospective neuropsychological assessment, and 2) retrospectively over the 48 months prior to study date. Neuropsychological assessment reports and patient electronic records will be screened to determine if participants potentially meet the inclusion and exclusion criteria. This screening will be conducted by a member of the healthcare team within the neuropsychology service. Sources of identifiable information are the referrals record for neuropsychological assessment, electronic patient record system, and neuropsychological assessment reports. Potential participants’ will only be approached by a member of the healthcare team initially. A comprehensive screening of inclusion and exclusion criteria will take place once potential participants consent to study inclusion. Participants will be advised to take a minimum of 24 hours to consider participation before giving consent. Participants may wish to take longer to decide, and the study will remain open to opt in to as long as they remain eligible and can complete the 14-week participation period within the timeframe of the study completion. Participants are not paid to participate in the study but will have access to a travel budget of £100 to compensate for travel expenses.

### 2.10 Patient and Public Involvement

#### 2.10.1 Pre-study PPI

The study has been informed by over 20 years of feedback from families affected by childhood brain tumour referred to the Neuropsychology service at GOSH. The intervention was initially developed for a patient and their carer at GOSH who reported favourable outcomes. Twelve families who had recently received care within the GOSH Neuropsychology service provided feedback on the funding proposal. The feedback was highly positive and indicated a high level of demand with 11 of 12 families indicating they would be interested in taking part. Most families said that they would find it beneficial to have options for online participation and/or support with travel costs, and for sessions to fit around school hours. All feedback has been incorporated into the study design and budget. The study was also informed by a recent North Thames survey (8) of forty-five families of childhood survivors of PBT where only 2% reported being able to access dedicated cognitive rehabilitation support, and 69% stating they either definitely would have liked their child to receive it (47%) or were unsure (22%).

The Participants Information Sheets (PISs) for the study were developed using guidance and templates from the Sponsor’s Patient and Public Involvement (PPI) department and GOSH Young Person Advisory Group (YPAG) of service users. The GOSH YPAG provided advice on the acceptability of the study design, use of plain English for study documents including the PIS, and recruitment practices. All advice from the YPAG has been incorporated into the study and protocol. A carer for someone treated for childhood brain tumour and healthcare service user provided further suggestions through NIHR Research for Patient Benefit (RfPB) peer review.

#### 2.10.2 PPI during the study

A Patient/Public Advisory Group (PAG) will be established at the start of the project and chaired by the PPI lead for the research study. The PAG has been appropriately costed for the study using NIHR payment guidance. A Stakeholder Mapping exercise under the Theory of Change model (44) was completed with the Chief Investigator and PPI study lead to identify stakeholders with relevant lived experience and knowledge of brain tumour diagnosis and treatment. The PAG will include 6 members. Nine 3-hour quarterly meetings will take place across the lifecycle of the study. The content of each PAG meeting will be dependent on the project stage. The group will be involved in providing advice and/or co-production for the study, including recruitment practices, creation of materials during the study, analysis, and dissemination of research.

## 3. ETHICS AND DISSEMINTATION

The study (Protocol number 22BO24 version 2, 16.12.2024) has been approved by the Camden & Kings Cross Research Ethics Committee (REC reference: 24/LO/0844, IRAS Project ID: 327316). The study is registered on ClinicalTrials.gov registry (ClinicalTrials.gov Identifier: NCT06770335, 13/01/2025: https://clinicaltrials.gov/study/NCT06770335).

### 3.1 Consent

Documented informed consent or assent will be sought by trained and delegated members of the research team for all participants after a discussion session based on the PIS. A copy of the PIS will be provided to participants. Special considerations for informed consent with children include PISs adapted for different ages (7-11 years, 12-15 years, and age 16+ years and a parent/carer version), assent forms for under age 16 (with parental consent) and consent form for 16+ years, and new PISs (and consent where appropriate), when children move into a new age bracket during the study. Participants will be informed that participation in the study is entirely voluntary, they can decline participation without giving a reason, they can withdraw at any time without giving a reason, and not taking part will have no consequences for their ongoing healthcare. Capacity will be presumed in the first instance (as stated in the Mental Capacity Act, 2005). For adolescent participants who can provide informed consent or where a parent consenting for their child loses capacity, the participant will not continue in the study and only de-identified data will be retained.

### 3.2 Confidentiality

All investigators and study site staff must comply with the requirements of the Data Protection Act 2018. A password protected Site Enrolment and Participant Log will be stored in duplicate on a secure GOSH Trust drive and encrypted external device and only accessible to members of the research team directly involved in recruitment, data quality control, and audit. The Site Enrolment and Participant Log will be stored in a separate location to the CRFs and other study data. Every participant will be allocated a unique Participant Identification Number (PID) on study entry. The PID consists of an unrelated random sequence of characters. The Site Enrolment and Participant Log will be the only document that will link the participants name and NHS number with the PID. At no point in presentations or publications of study data will individual patients be identified. Any direct quotes used will be anonymised and will not contain potentially identifiable information. Personal data will be stored for no longer than 12 months after study completion and will only be accessed where this is essential to the study.

### 3.3 Study monitoring

The Chief Investigator will be responsible for the day-to-day management of the study with support from the study management group. The study is a low risk non-CTIMP and does not require a data monitoring committee (DMC). A Trial Steering Committee (TSC) has been established to provide overall supervision for the study on behalf of the study’s Sponsor and Funder and to ensure that it is conducted to the rigorous standards set out in the UK Policy Framework for Health and Social Care and the Guidelines for Good Clinical Practice. The conduct of the research will be subject to monitoring and auditing according to the Joint Research & Development Office for GOSH/University College London - Institute of Child Health standard research operating procedures.

### 3.4 Risk and safety monitoring and reporting

The NIHR ‘Decision Tree for Adverse Event Reporting – Non-CTIMPS’, including standard definitions of an Adverse Event (AE) and Serious Adverse Event (SAE) will guide study monitoring. It is highly unlikely that a serious adverse event/reaction would be attributable to this low-risk non-CTIMP trial. However, all AEs and SAEs will be documented in an Adverse Event Reporting log and CRF. Where an SAE occurs, this will be reported to the Sponsor within 24 hours. Participants will be notified of relevant events within 7 days or prior to their next study contact if this is sooner. If during any study activity a participant or their carer discloses risk or safeguarding concerns (i.e., their intention to harm themselves or others), the local GOSH Trust risk assessment and safeguarding policy will be immediately followed.

### 3.5 Dissemination

The NIHR and Success Charity will be acknowledged in any dissemination. A full report of the study findings will be submitted for publication within 2 years of study completion. The formal findings will be submitted to peer-reviewed scientific journals and, where possible, presented at relevant conferences and shared with relevant stakeholders. A lay summary of the findings will be co-produced with our PAG for dissemination amongst service-users, charities, and relevant social media. We aim to use the findings from this project to apply for funding for a multicentre RCT. Participants who request results from the CI will be provided with this information after the results of the study have been published.

Authors will be individually named on the final study report. Authors will have made a substantive intellectual contribution to the report and guided by The International Committee of Medical Journal Editors recommended authorship criteria: https://www.icmje.org/recommendations/browse/roles-and-responsibilities/defining-the-role-of-authors-and-contributors.html.

## 4. CONCLUSIONS

There is a stark discrepancy between the high level of cognitive morbidity for childhood survivors of brain tumour and available rehabilitative support. An essential first step is to better understand feasibility and acceptability of cognitive rehabilitation for this population, particularly with reference to highly prevalent co-morbidities such as cognitive fatigue. Efficacy measures should clearly map to the aims and purpose of the interventions and play an iterative role in the development of theoretical frameworks for developing and trialling rehabilitation interventions. As is the case in many areas of paediatric acquired brain injury, we are in the early stages of understanding if and how cognitive rehabilitation interventions could work for this population and how this may be implemented within the NHS. The current study aims to meet a substantial evidence gap and answer these essential questions for this vulnerable population of children.

## Data Availability

No data are associated with this article (protocol). The SPIRIT checklist for the study protocol is accessible on the Figshare repository

https://doi.org/10.6084/m9.figshare.28225142.v1

## Data (and Software) Availability

No data are associated with this article.

## Reporting guidelines

The study protocol and protocol manuscript were written in line with the SPIRIT 2013 checklist. The SPIRIT checklist for the study protocol is accessible on the Figshare repository: https://doi.org/10.6084/m9.figshare.28225142.v1

## Award Information

This project is funded by the National Institute for Health and Care Research (NIHR) under its Research for Patient Benefit (RfPB) Programme (Grant Reference Number NIHR208009). The views expressed are those of the authors and not necessarily those of the NIHR or the Department of Health and Social Care.

The patient involvement elements of this study are funded by Success Charity Life After Cure Limited (SCLAC, Grant Reference Number SUCCESS002) as part of its mission to transform recovery care from a childhood brain tumour, alongside cure. The views expressed are those of the authors and not necessarily those of SCLAC.

## Acknowledgements

We thank Dr Sophie Thomas for the review and advice provided on the initial research plan.

